# The distribution of regions of homozygosity (ROH) among consanguineous populations - implications for a routine genetic counseling service

**DOI:** 10.1101/2024.05.07.24306935

**Authors:** Chen Gafni-Amsalem, Nasim Warwar, Morad Khayat, Yasmin Tatour, Olfat Abuleil-Zuabi, Salvatore Campisi-Pinto, Shai Carmi, Stavit A. Shalev

**Author notes:** **Corresponding author:** Chen Gafni-Amsalem, Genetics Institute Emek Medical Center, Afula 18101, Israel., Phone: work 972-4-649-5446.

## Abstract

**Background:** Regions of homozygosity (ROH) increase the risk of recessive disorders, and guidelines recommend reporting of excessive ROH in prenatal testing. However, ROH are common in populations that practice endogamy or consanguinity, and cutoffs for reporting ROH in such populations may not be evidence-based.

**Methods:** We reviewed prenatal testing results (based on cytogenetic microarrays) from 2191 pregnancies in the Jewish and non-Jewish populations of Northern Israel and estimated the prevalence of ROH according to self-reported ethnicity and parental relationships.

**Results:** The proportion of the genome in ROH, ROH rate, was higher in non-Jews [Mean(SD)=2.91%(3.92%); max=25.54%; N=689] than in Jews [Mean(SD)=0.81%(0.49%); max=3.93%; N=1502]. In the non-Jewsih populations, consanguineous marriages had the highest ROH rates [Mean(SD)=7.14%(4.55%), N=217], followed by endogamous [Mean(SD)=1.13%(1.09%), N=283]) and non-endogamous [Mean(SD)=0.69%(0. 56%), N=189]) marriages. ROH rates were greater than 5%, the ACMG-recommended cutoff, in 149/689 (21.63%) of the non-Jewish samples. Within the Jewish populations, the rates were similar between Ashkenazi, North African, and Middle Eastern Jews, but were higher for six consanguineous unions [Mean(SD)=2.38%(1.23%)] and when spouses belonged to the same sub-population.

**Conclusions:** Given the high ROH rates we observed in some subjects, we suggest that assessing the risk for recessive conditions in consanguineous/endogamous populations should be done before the first pregnancy, through genetic counseling and sequencing. Such an approach will: (1) identify couples who are at risk and counsel them on reproductive options; and (2) avoid the stress that couples who are not at risk may experience due to a prenatal ROH report.

## Introduction

The process of detecting regions of homozygosity (ROH) through chromosomal microarray (CMA) is presently considered standard practice for both prenatal and postnatal specimens, rather than an incidental finding (1). The plausible mechanisms for the presence of ROH are diverse and may include population history events such as bottleneck or geographic isolation; as well as cultural practices that promote consanguineous marriage and endogamy (2). In this scenario, the chromosomal segments in ROH are inherited from a common ancestor of the parents, and are thus identical by descent (IBD) (3,4). Another mechanism behind ROH may reflect Uniparental Disomy (UPD). UPD occurs when both copies of a chromosome, or segments of it, are inherited from one parent (in the absence of the chromosome from the other parent). Among the possible clinical consequences are conditions associated with abnormal imprinting processes, in case the ROH segment contains imprinted genes (5).

Consanguinity confers increased genome homozygosity, leading to a higher risk of autosomal recessive disorders (6). Therefore, the presence of ROH may require the use of additional genetic tools to rule out recessive diseases, such as exome or genome sequencing (7, 8).

The coefficient of inbreeding (F) gives the average proportion of the offspring’s autosomal genome that is inherited from a common ancestor. Theoretically, individuals who are offspring of first cousins or double-first cousin mating should present ROH of (F=1/16) 6.25% and (F=1/8) 12.5% of their genome, respectively (9). However, significant deviations from the expected values may occur due to several reasons, including random Mendelian segregation of chromosomes during meiosis, families with multiple loops of consanguinity, or endogamous communities (multiple generations of breeding within a relatively closed community) (6,10).

The American College of Medical Genetics and Genomics (ACMG) has established technical standards for interpreting and reporting regions of homozygosity and suspected consanguinity or UPD (1). The recommended cutoff was ROH <5% of the autosomal genome for fetal testing via prenatal CMA to cover most consanguineous close mating cases (1). Such reports require elaboration of the ramifications of the finding through genetic counseling process, adding to the workload of healthcare professionals in genetics.

Long ROH segments may indicate recent parental relatedness. Such segments were found to be more frequent and variable in non-Jewish populations from the Middle East and Central/South Asia, and of several widely dispersed Jewish populations (2,9). A previous study of the Druze communities (from Galilee, Golan, Carmel in Israel, and from Lebanon) found a large proportion of individuals with multiple long ROH (11).

The Israeli population, as of 2023, comprises 9.795 million people, of whom 7.181 million (73%) are Jews, 2.065 million (21%) Arabs, and 549,000 (6%) others (12). These various ethnic groups each has its own demographic history. Each sub-group is characterized by an increased frequency of specific heredity disorders (13).

The non-Jewish Arab population is often found in small and isolated villages/small towns. The preference for consanguineous marriages in some Arab communities yields an increased risk of autosomal recessive disorders in their progeny (6). Consanguineous marriages are becoming less prevalent among the Arab populations in the Middle East, but they remain considerably high. It has been reported that endogamy occurs at a rate exceeding 70% (13). In such populations, rare recessive diseases are expected to occur more frequently due to the increase of homozygosity (6).

The Israeli Jews are mainly urban, and are roughly divided into Ashkenazi (European origin) and non-Ashkenazi. The Ashkenazi Jewish population, during its existence in Central and Eastern Europe, maintained genetic isolation from their neighbors due to religious and cultural practices (14). Such isolation is reflected by the high prevalence of autosomal-recessive diseases and the relatively high frequency of alleles that increase the risk of common disorders such as breast and ovarian cancer (15).

The Genetic Institute at Emek Medical Center, Afula, serves a large Arab population living in the North of Israel, mostly in the Jezreel Valley, lower Galilee, and the Eron Valley (Wadi Ara) (6). The Arab population of Northern Israel usually consists of large, multigenerational families with multiple loops of consanguinity (16). Among the Arabic population in North Israel, the consanguinity rate was documented in 2015 as 24.4% (17). It has been observed that the rate of malformations and genetic diseases among children born to consanguineous couples in North Israel is 6.8% (6).

This research aims to estimate and characterize the distribution of rates of ROH among different subgroups in North Israel, broken down by non-endogamous, endogamous, and consanguineous mating practices, hence, helping to establish standards of care including recommendations regarding genetic counseling.

## Methods

The data analyzed in the study included retrospectively reviewed cytogenetic microarray (CMA) results of 2191 prenatal tests between January 2020 and May 2023 and self-report information regarding the ethnicity of the parents and their familial relationships. Out of 120 women who had more than one chromosomal test, eleven had twin pregnancies, and ROH was computed for each co-twin separately. The CMA tests were performed according to medical indications of current national guidelines, including advanced maternal age, abnormal screening tests for Down syndrome, abnormal sonographic findings during pregnancy (including skeletal abnormalities, high nuchal translucency, anomalies of the brain, cardiovascular system, gastrointestinal tract, and other anomalies), suspected viral infection, consuming teratogenic drugs in pregnancy, molecular diagnosis of familiar monogenic disease, or according to maternal request.

The CMA analyses were done using Affymetrix® - Thermo Fisher Scientific Inc. (Life Technologies, Carlsbad, CA, USA). The data was analyzed, including the detection of ROH, using Affymetrix Chromosome Analysis Suite software (ChAS – Santa Clara, CA, USA) version 4.4.1, with 200,436 single nucleotide polymorphism (SNP) markers. The ROH reporting threshold was >3Mb, with a minimum of 50 markers. SNPs on the sex chromosomes were excluded.

The percentage of homozygosity (%ROH) in the genome of each individual was calculated by dividing the sum of all homozygous regions in the autosome by the total autosomal length (2780Mb for GRCh37 – hg19) and multiplying the result by 100 (10).

The expected coefficient of inbreeding (*F*) was based on the degree of the biological reported relationship between the parents, including double first cousins (*F*=1/8), first cousins (*F*=1/16), first cousin once removed or ‘distant’ (*F*=1/32), and unrelated (*F*=0) (9, 18). We classified the non-Jewish populations into three major groups: Non-endogamous (unrelated spouses who were also born in different villages/towns); Endogamous (unrelated spouses who originated from the same village; and/or individuals of Christian Arab origin, who mostly have Syrian-Lebanese origin); and Consanguineous, defined according to the obtained pedigree.

The Jewish population was grouped into Ashkenazi, Middle Eastern (including origins in Iran, Iraq, Kurdistan, Yemen, Caucasus, Egypt, Turkey, and Syria), North African (including Morocco and Tunisia), and Ethiopian. “Other mixed origins” include combinations of two origins or more. In pregnancies designated as mixed origins, each of the parents can be themselves of mixed origins.

This study was approved by the Ethical Committee of Emek Medical Center (EMC-0073-23), following the Helsinki guidance.

Statistical analysis was performed using IBM SPSS version 28.0.1.1 and Excel computer programs. Descriptive statistics were performed for the measured parameters. Outliers are represented in all graphs by circles (for values that are more than 1.5xIQR below Q1 or above Q3); or by asterisks (for values that are more than 3.0xIQR below Q1 or above Q3. Differences between groups were examined by the one-sample t-test, the Mann-Whitney test, and the Kruskal-Wallis rank sum tests. P<0.05 was considered statistically significant. Post hoc estimates were corrected using the Bonferroni method.

## Results

This study included CMA tests that were performed in 2191 pregnancies. The observed ROH rates were in the narrow range from 0 to an upper quartile of 1.27%, but with a maximum of 25.54%. The ROH rate was significantly higher among non-Jews vs. Jews (U =-678374.50, P<0.001), see Figure 1, Table 1.

**Figure 1:**
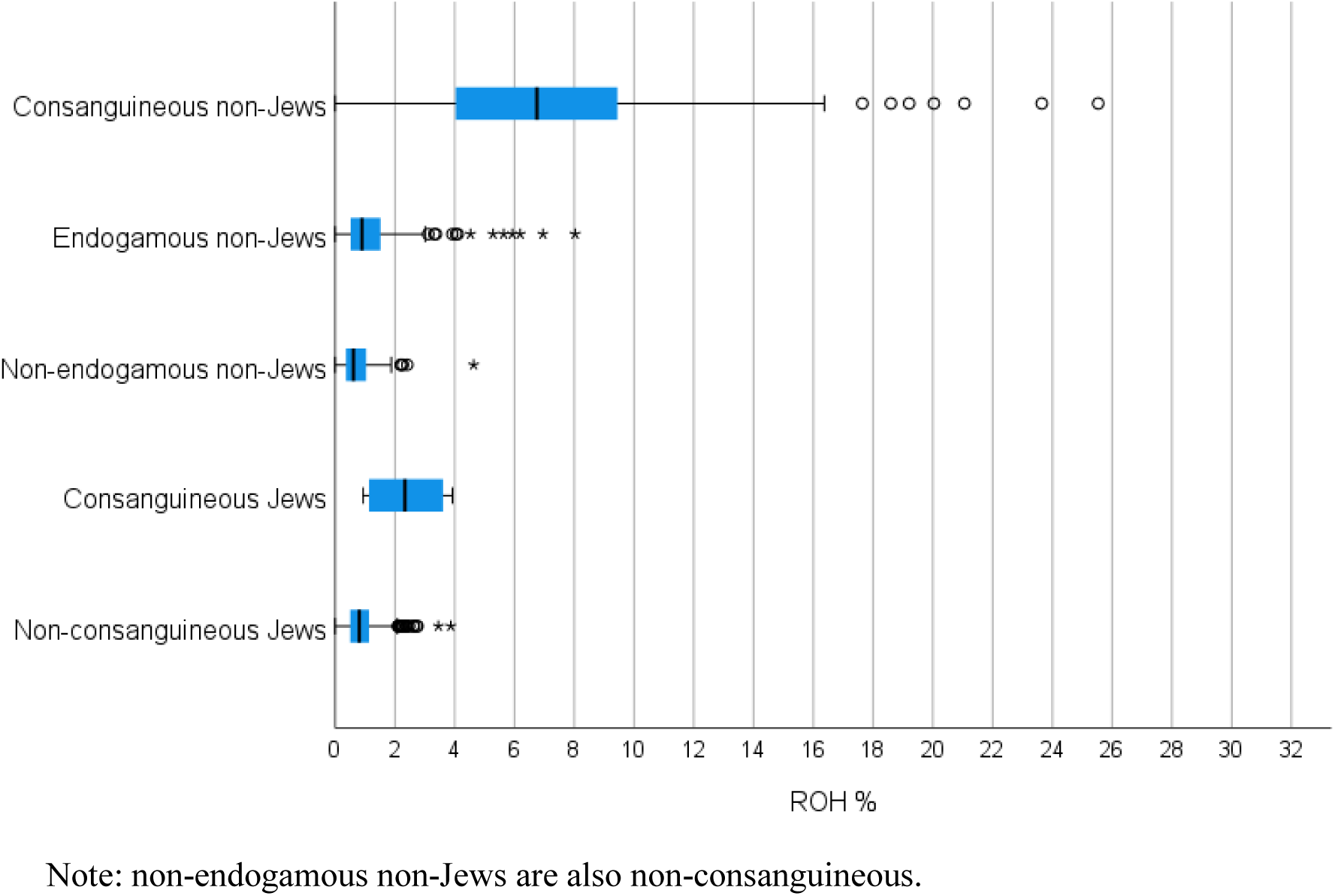
ROH rates in Jewish and non-Jewish groups who practice consanguineous, endogamous, and non-endogamous mating.

**Table 1:**
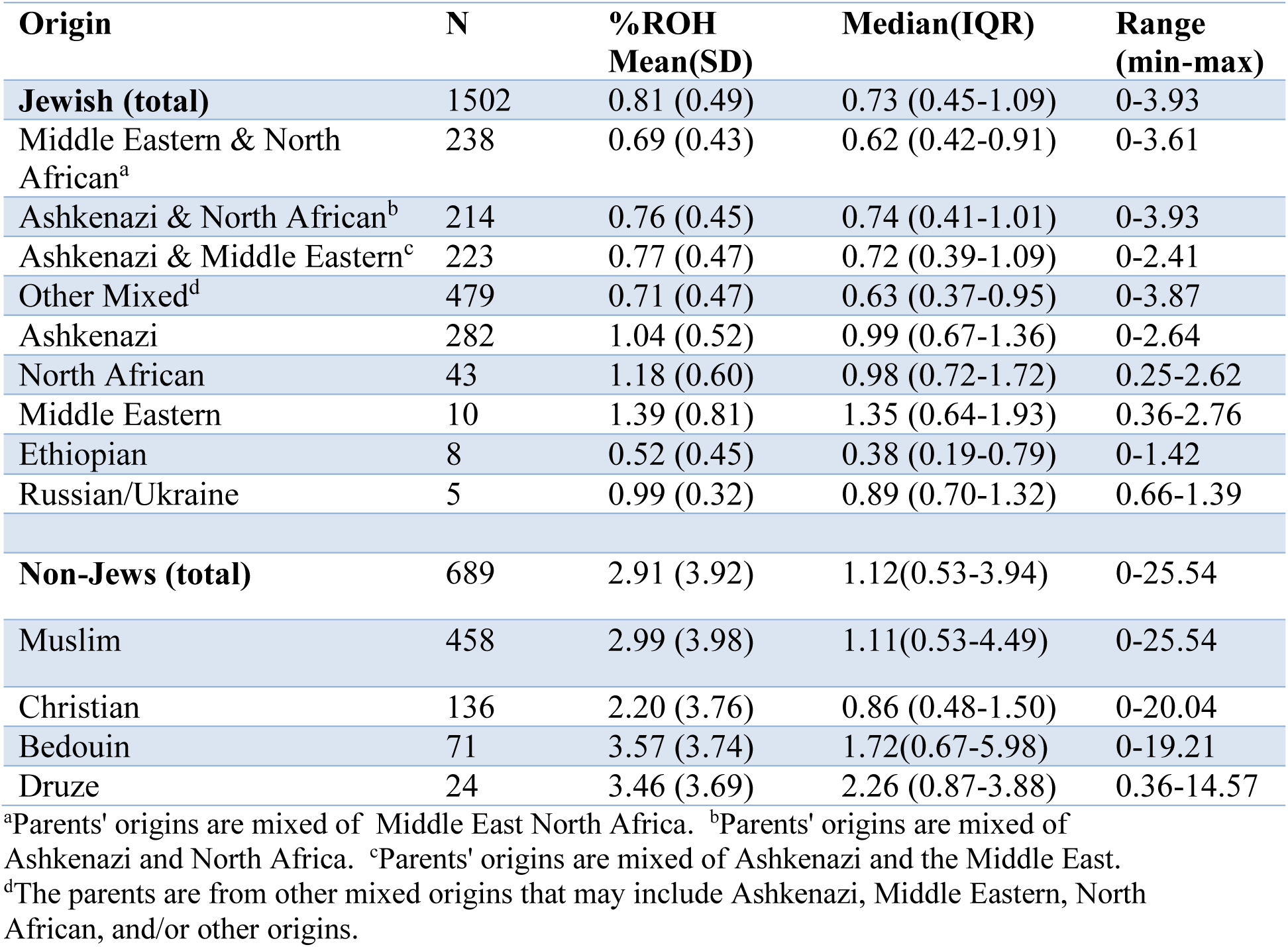
ROH rate according to parents’ ethnic group.

| Origin | N | %ROH<br>Mean(SD) | Median(IQR) | Range<br>(min-max) |
| --- | --- | --- | --- | --- |
| **Jewish (total)** | 1502 | 0.81 (0.49) | 0.73 (0.45-1.09) | 0-3.93 |
| Middle Eastern & North African<sup>a</sup> | 238 | 0.69 (0.43) | 0.62 (0.42-0.91) | 0-3.61 |
| Ashkenazi & North African<sup>b</sup> | 214 | 0.76 (0.45) | 0.74 (0.41-1.01) | 0-3.93 |
| Ashkenazi & Middle Eastern<sup>c</sup> | 223 | 0.77 (0.47) | 0.72 (0.39-1.09) | 0-2.41 |
| Other Mixed<sup>d</sup> | 479 | 0.71 (0.47) | 0.63 (0.37-0.95) | 0-3.87 |
| Ashkenazi | 282 | 1.04 (0.52) | 0.99 (0.67-1.36) | 0-2.64 |
| North African | 43 | 1.18 (0.60) | 0.98 (0.72-1.72) | 0.25-2.62 |
| Middle Eastern | 10 | 1.39 (0.81) | 1.35 (0.64-1.93) | 0.36-2.76 |
| Ethiopian | 8 | 0.52 (0.45) | 0.38 (0.19-0.79) | 0-1.42 |
| Russian/Ukraine | 5 | 0.99 (0.32) | 0.89 (0.70-1.32) | 0.66-1.39 |
| **Non-Jews (total)** | 689 | 2.91 (3.92) | 1.12(0.53-3.94) | 0-25.54 |
| Muslim | 458 | 2.99 (3.98) | 1.11(0.53-4.49) | 0-25.54 |
| Christian | 136 | 2.20 (3.76) | 0.86 (0.48-1.50) | 0-20.04 |
| Bedouin | 71 | 3.57 (3.74) | 1.72(0.67-5.98) | 0-19.21 |
| Druze | 24 | 3.46 (3.69) | 2.26 (0.87-3.88) | 0.36-14.57 |

<sup>a</sup>Parents' origins are mixed of Middle East North Africa. <sup>b</sup>Parents' origins are mixed of Ashkenazi and North Africa. <sup>c</sup>Parents' origins are mixed of Ashkenazi and the Middle East.
<sup>d</sup>The parents are from other mixed origins that may include Ashkenazi, Middle Eastern, North African, and/or other origins.

### Non-Jewish populations

Among the non-Jewish sub-populations, the ROH rate was significantly lower among Christian Arabs compared to Bedouin (H=108.265, P_adj_=0.001), and Druze (H=-124.437, P_adj_=0.028), but not compared to non-Bedouin Muslim Arabs (H=-47.683, P_adj_=0.085), Table 1.

There was no significant difference in ROH rate between non-Bedouin Muslim Arabs and Bedouins (H=60.582, P_adj_=0.102), non-Bedouin Muslim Arabs and Druze (H=76.754, P_adj_=0.393), or Druze and Bedouins (H=-16.173, P_adj_=1.000), Table 1.

Within the non-Jewish populations, consanguineous mating had the highest ROH rate [Median(IQR)=6.69(3.79-9.37), Mean(SD)=7.14(4.55), N=217] compared to endogamous [Median(IQR)=0.86(0.51-1.37), Mean(SD)=1.13(1.09), N=283; H=281.545, P_adj_<0.001] and non-endogamous mating [Median(IQR)=0.57(0.32-0.89), Mean(SD)=0.69(0.56), N=189; H=358.414, P_adj_<0.001], Figure 1. The ROH rate was significantly higher for endogamous vs. non-endogamous mating (H=76.868, P_adj_<0.001).

In 149 out of 2191 of the individuals (6.80%), all belonging to the non-Jewish populations (149/689, 21.63%), the ROH rates were greater than 5%. In 143 out of the 149 (96%), the parents were relatives, and the rest were endogamous couples.

### Consanguinity

A higher ROH rate was noticeably more prevalent among fetuses of consanguineous versus non-consanguineous spouses (Figure 1). Rates were higher among consanguineous non-Jews [Median(IQR)=6.69(3.79-9.37), Mean(SD)=7.14(4.55), N=217] vs. consanguineous Jews [Median(IQR)=2.34(1.08-3.69), Mean(SD)=2.38(1.23), N=6], although sample size is too small for hypothesis testing.

Non-consanguineous Jews [Median(IQR)=0.73(0.45-1.08), Mean(SD)=0.79(0.48), N=1496] had higher ROH rates than non-consanguineous and non-endogamous non-Jews [Median(IQR)=0.57(0.32-0.89), Mean(SD)=0.69(0.56), N=189], U=117692.0, p<0.001.

Among the 223 consanguineous cases (out of 2191, 10.18%), 6 couples were Jewish, 34 were Bedouin, 6 were Druze, 153 were Muslim Arabs, and 24 were Christian Arabs.

In Figure 2 and Table 2, we show the ROH rates per self-reported parental relationship for the consanguineous non-Jewish families. The mean/median observed ROH rates were higher than expected for most types of relationships. Of note, one Muslim-Arab consanguineous couple (I+II cousins) had dizygotic twin pregnancy with one fetus having ROH of 4.56%, and the other 10.14%. Ten additional dizygotic twin pregnancies were documented with more similar rates between the co-twins.

**Figure 2:**
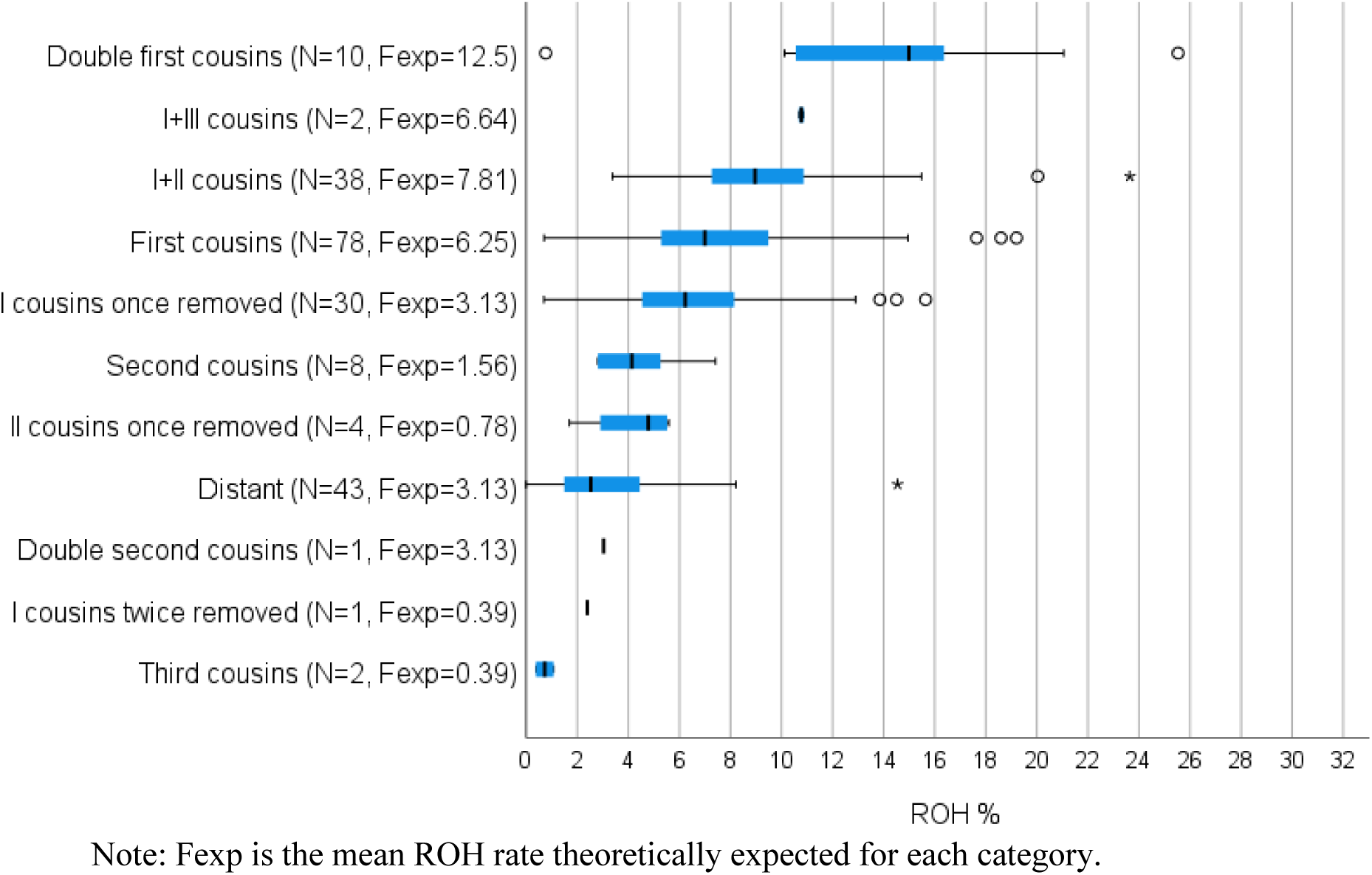
ROH rates among consanguineous non-Jews, according to the relationship between the parents.

**Table 2:** ROH rates among consanguineous non-Jews in descending order of expected ROH percentage.

| *Consanguinity* | *N* | *%ROH*<br>*mean(SD)* | *Median(IQR)* | *Range*<br>*(min-max)* | *ExpF<sup>a</sup> (%)* | *Statistics<sup>b</sup>* |
| --- | --- | --- | --- | --- | --- | --- |
| **Double first cousins** | 10 | 14.35(6.65) | 14.99(10.45-17.54) | 0.76-25.54 | 12.5 | t=0.882<br>P=0.401 |
| **I+III cousins** | 2 | 10.78(0.13) | 10.78(10.69-10.78) | 10.69-10.88 | 6.64 | - |
| **I+II cousins** | 38 | 9.61 (4.02) | 8.97(7.27-11.04) | 3.37-23.65 | 7.81 | t=2.764<br>P=0.009 |
| **First cousins** | 78 | 7.71(3.79) | 6.99(5.28-9.49) | 0.70-19.21 | 6.25 | t=3.393<br>P<0.001 |
| **I cousins once removed** | 30 | 6.98(3.69) | 6.23(4.47-8.44) | 0.69-15.64 | 3.13 | t=5.724<br>P<0.001 |
| **Second cousins** | 8 | 4.33(1.63) | 4.14(2.81-5.31) | 2.80-7.42 | 1.56 | t=4.825<br>P=0.002 |
| **II cousins once removed** | 4 | 4.22(1.81) | 4.79(2.29-5.56) | 1.69-5.59 | 0.78 | t=3.790<br>P=0.032 |
| **Distant** | 43 | 3.50(2.85) | 2.53(1.49-4.52) | 0-14.55 | 3.13 | t=0.862<br>P=0.393 |
| **Double second cousins** | 1 | 3.03 |  | - | 3.13 | - |
| **I cousins twice removed** | 1 | 2.39 |  | - | 0.39 | - |
| **Third cousins** | 2 | 0.73(0.49) | 0.73(0.38-0.73) | 0.38-1.08 | 0.39 | - |

<sup>a</sup> ExpF is the expected value (in percentage) of the coefficient of inbreeding, F.
<sup>b</sup> Comparison of observed ROH percentage to the expected F, according to the one-sample t-test.

The six consanguineous Jewish couples reported various relationships, including two couples who are second cousins once removed, three couples second cousins, and one couple who were third cousins.

### ROH among Jewish populations

ROH rate among fetuses of Jewish spouses was in the range of 0 to 3.93%, with lower values among admixed couples and Ethiopian Jews (Table 1, Figure 3).

**Figure 3:**
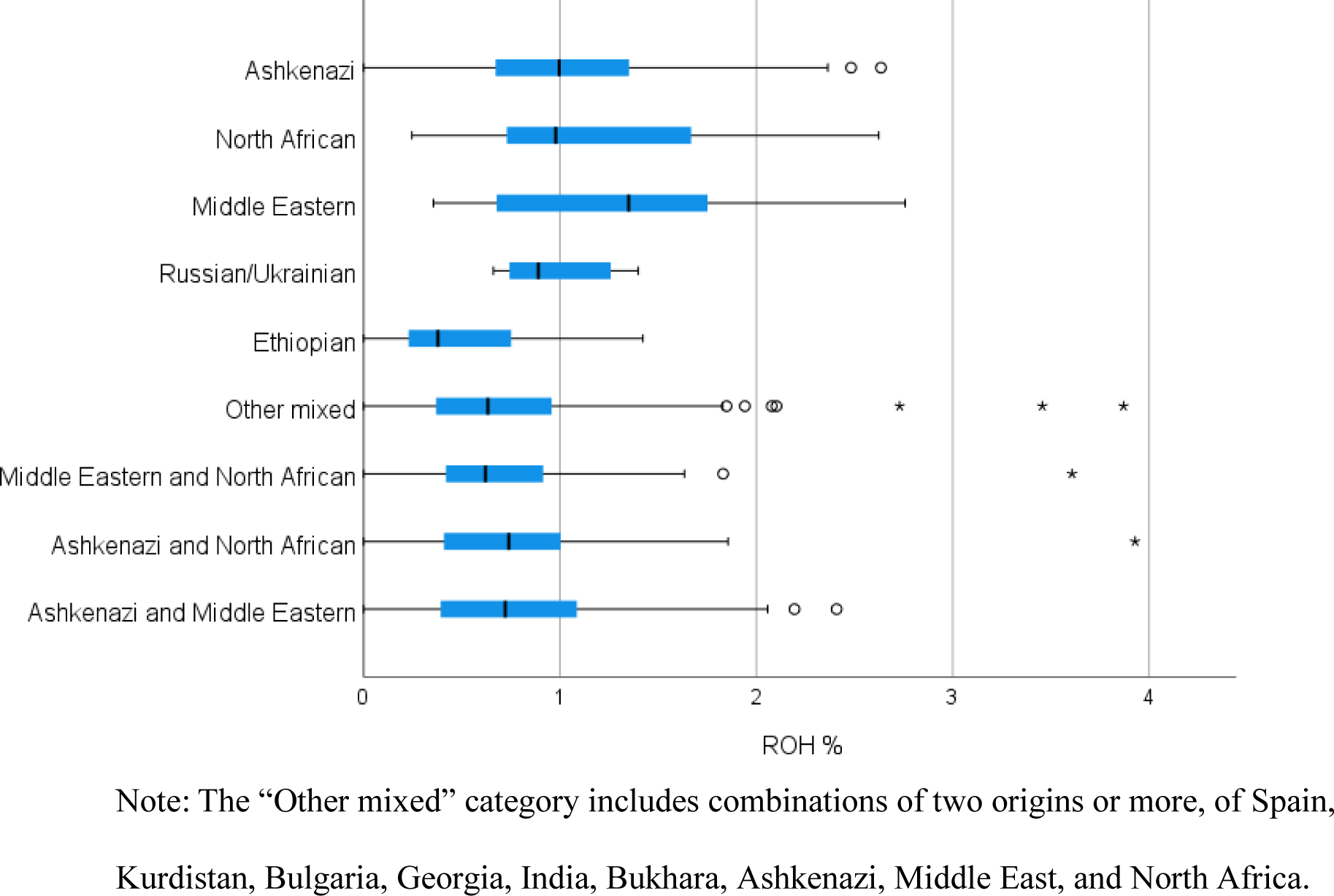
ROH rates among Jews according to ethnic origin.

Ashkenazi Jews had a similar ROH rate to that of North African Jews (H=-69.232, P_adj_=1.000) and Middle Eastern Jews (H=-130.174, P=1.000) (Table 1, Figure 3).

Ashkenazi Jews had higher ROH values compared to all Jews with mixed origins (P_adj_<0.001 for all comparisons; Table 1). North African Jews also had higher ROH values compared to all Jews with mixed origins (P_adj_<=0.001 for all comparisons; Table 1). The groups with mixed Jewish origins had no significant difference in ROH rate between each other (P_adj_=1.000 for all comparisons).

Ethiopian Jews had significantly lower ROH values than North African Jews (P_adj_=0.034). Other comparisons involving Jewish groups had no significant differences in ROH rate (P_adj_>=0.068), Table 1.

## Discussion

This study presents the ROH rates as calculated on CMA analyses of 2191 pregnancies of various Israeli sub-populations. Since the risk for uniparental disomy (UPD) is not expected to differ across populations, we henceforth focus only on the consequences of abnormal length of ROH to the risk for autosomal recessive conditions. The average percentage of ROH among Jews and non-Jews in our cohort was 0.81% and 2.91%, respectively. The distribution of ROH rates was right-tailed in the non-Jewish population, with a median of 1.12%, but the range reached 25.54%.

When focusing on consanguineous marriages, there is a striking difference between Jews to non-Jews: for the six Jewish couples reported to be related to each other, the median ROH was 2.34% and the maximum reached 3.93%. Among the non-Jewish population, the picture was different - for most reported types of relationship between couples, the observed mean or median ROH was significantly higher than expected according to theoretical values, sometimes by a large margin, indicating that most of these couples are more related to each other than they knew or reported, an observation we previously made (6). This included several cases where parents self-reported as second cousins, or “distant” familial relations, but the ROH rate exceeded 5%. In five fetuses, the ROH rates were close to those expected when the parents were first-degree relatives (19-25%).

According to the ACMG, the recommended cutoff for reporting ROH of prenatal CMA was >5% of the autosomal genome, to cover most consanguineous close mating cases (1). In our cohort, 149 out of the 689 non-Jews (21.63%) presented ROH greater than 5%, most of them were relatives. None of the 1502 Jewish samples had ROH>5%. Given our results, it is clear that the most appropriate timing to assess the risk for autosomal recessive conditions in consanguineous/endogamous populations appears to be prior to the first pregnancy, with a structured genetic counseling process and next-generation sequencing of the future parents. This is expected to be more effective and more appropriate in terms of resources use compared to the prenatal stage. In addition, counseling these couples during pregnancies will dramatically contribute to their chaotic emotional state. The clinical benefit of such a recommendation fits the unique features of the Israeli Arab population, including a high fertility rate, underutilization of prenatal diagnosis services (16), and low rate of pregnancy termination of an affected fetus (16).

Endogamous non-Jewish populations demonstrated higher ROH rates compared to non-endogamous, and lower than consanguineous unions. Although overall, the mean/median rate of ROH has not exceeded 5% in this sub-group, still some samples presented higher rates reaching 8% of the genome. Therefore, it seems that couples who were both born in the same town/village must all be regarded as at high risk for autosomal recessive conditions. Having preliminary knowledge about the more frequent autosomal recessive variants in any of these cohorts might benefit future couples. Exome sequencing of 50 random, healthy adults from a single Muslim-Arab village in northern Israel revealed 48 autosomal recessive variants, of which 24 variants had not been previously detected in this population (19). It emphasizes the importance of preconception and personalized genetic counseling in this population rather than prenatal testing.

Within the Jewish populations, the ROH rate was higher when the spouses shared the same ancestral origin compared to mixed origins, as expected. Similar ROH rates were documented among Ashkenazi, North African, and Middle Eastern Jews. In contrast to our results, Kang et al. (9) and Waldman et al (20) found that the ROH values of Jewish populations from the Middle East, North Africa, and South Asia are generally higher than in Ashkenazi populations. These differences, which require further exploration, may reflect the limited sample size in these studies or differences between generations, as consanguinity practices can change over time. The Ethiopian Jewish community has a longstanding cultural practice of eschewing consanguineous marriages. To this end, they maintain familial ancestral records for up to seven generations (21), a tradition that is associated with the low ROH rate found by Kang et al. (9) and in our study.

In conclusion, we suggest that the ACMG recommendation of reporting ROH greater than 5% in prenatal testing should not apply to populations with a long tradition of endogamy and/or current consanguineous marriages. In these populations, counseling future parents prior to their first pregnancy is expected to be more beneficial, given that a large fraction of children will be born with ROH. This will serve two purposes. The first is to determine whether a couple is at risk of a recessive disease prior to a pregnancy, and then provide more informed counseling regarding their reproductive options. Second, avoiding the stress to couples when receiving a result of a possible genetic abnormality during prenatal testing. Given that a large fraction (21.63%%) of our non-Jewish subjects displayed ROH rates >5%, reporting such ROH may lead to more harm than benefits (22). This underscores the importance of programmed genetic counseling and using panel or exome sequencing to identify pathogenic autosomal recessive variants in couples from these populations.

## Data Availability

All data produced in the present study are available upon reasonable request to the authors

## Author contribution

Conceptualization: CGA, NV, MK, YT, SAS; formal analysis: CGA, SCP; methodology: CGA, NV, YT, OAZ, SC, SAS; investigating: CGA, NV, YT, SC, SAS; project administration: CGA, NV, MK, YT, OAZ, SAS; supervision: SAS; validation: CGA, SC, OAZ, NV, MC, SAS; writing—original draft: CGA; writing, review and editing: CGA, SC, SAS.

## Ethics declarations

The study protocol was approved by the ethical committee of the Emek Medical Center, Afula, following the Helsinki Declaration. Research number EMC-0073-23. All procedures followed were in accordance with the ethical standards of the responsible committee on human experimentation (institutional and national) and with the Helsinki Declaration of 1975, as revised in 2000.

